# Point of Care Testing using rapid automated Antigen Testing for SARS-COV-2 in Care Homes – an exploratory safety, usability and diagnostic agreement evaluation

**DOI:** 10.1101/2021.04.22.21255948

**Authors:** Massimo Micocci, Peter Buckle, Gail Hayward, A. Joy Allen, Kerrie Davies, Patrick Kierkegaard, Karen Spilsbury, Carl Thompson, Anita Astle, Ros Heath, Claire Sharpe, Cyd Akrill, Dan Lasserson, Rafael Perera, Richard Body, Adam L Gordon

**Affiliations:** NIHR London In Vitro Diagnostics Co-operative, Department of Surgery and Cancer, Imperial College London, London, UK; NIHR Community Healthcare Medtech and IVD cooperative, Nuffield Department of Primary Care Health Sciences, University of Oxford, UK; NIHR Newcastle In Vitro Diagnostics Co-operative, Translational and Clinical Research Institute, Newcastle University, Newcastle Upon Tyne, UK; Healthcare Associated Infections Research Group, University of Leeds and Leeds Teaching Hospitals NHS Trust, Leeds, UK; NIHR Leeds In Vitro Diagnostics Co-operative, Leeds Teaching Hospitals NHS Trust, Leeds, UK; CRUK Convergence Science Centre, Institute for Cancer Research & Imperial College London, London, UK; School of Healthcare, University of Leeds, Leeds, UK; NIHR Applied Research Collaboration Yorkshire and Humber, UK; Wren Hall Nursing Home, Selston, UK; Landermeads Nursing Home, Nottingham, UK; Ashmere Nottinghamshire Ltd, Notts, UK; Springfield Healthcare, Leeds, UK; Warwick Medical School, University of Warwick, Coventry, UK; Nuffield Department of Primary Care Health Sciences, University of Oxford, Oxford, UK; NIHR Applied Research Collaboration - Yorkshire and Humber (YHARC), UK; Division of Medical Sciences and Graduate Entry Medicine, School of Medicine, University of Nottingham, UK; NIHR Applied Research Collaboration-East Midlands (ARC-EM), Nottingham, UK

## Abstract

**Introduction:** Successful adoption of POCTs (Point-of-Care tests) for COVID-19 in care homes requires the identification of ideal use cases and a full understanding of contextual and usability factors that affect test results and minimise biosafety risks. This paper presents findings from a scoping-usability and test performance study of a microfluidic immunofluorescence assay for COVID-19 in care homes.

**Methods:** A mixed-methods evaluation was conducted in four UK care homes to scope usability and to assess the agreement with qRT-PCR. A dry run with luminescent dye was carried out to explore biosafety issues.

**Results:** The agreement analysis was carried out on 227 asymptomatic participants (159 staff and 68 residents) and 14 symptomatic participants (5 staff and 9 residents). Asymptomatic specimens showed 50% (95% CI: 1.3%-98.7%) positive agreement and 96% (95% CI: 92.5%-98.1%) negative agreement with overall prevalence and bias-adjusted Kappa (PABAK) of 0.911 (95% CI: 0.857-0.965). Symptomatic specimens showed 83.3% (95% CI: 35.9%-99.6%) positive agreement and 100% (95% CI: 63.1%-100%) negative agreement with overall prevalence and bias-adjusted Kappa (PABAK) of 0.857 (95% CI: 0.549-1).

The dry run showed four main sources of contamination that led to the modification of the standard operating procedures. Simulation after modification showed no further evidence of contamination.

**Conclusion:** Careful consideration of biosafety issues and contextual factors associated with care home are mandatory for safe use the POCT. Whilst POCT may have some utility for ruling out COVID-19, further diagnostic accuracy evaluations are needed to promote effective adoption.

## Background

Protecting care homes from COVID-19 is complex[1] [2] [3] and will continue to be challenged by immunosenescence in care home populations [4] and the emergence of new strains [5], even after the widespread roll-out of vaccines. Minimising ingress into homes [6] can help reduce the risk of infection but restricting visiting can cause physical and mental deconditioning in residents and may contravene fundamental human rights [7].

Testing regimes may help facilitate safe opening of care homes by enabling early identification of infection through rapid establishment of COVID-19 status for those who are symptomatic, and through regular screening of those who are asymptomatic. Laboratory based reverse-transcriptase polymerase chain reaction (RT-PCR) tests are considered to be highly sensitive and specific but results may take more than 24 hours to be received, and sometimes several days to return to care homes, making them suboptimal for outbreak management [8]. Widely deployed lateral flow device (LFD) antigen tests provide results within 30 minutes but normally have lower sensitivity and specificity than RT-PCR and present other use challenges in a care home context [9, 10]. Changes in sensitivity of LFDs have been reported as a direct consequence of the context of use and when carried out by untrained healthcare workers [11, 12]. These findings demonstrate that successful adoption of a diagnostic test requires a fuller understanding of all the factors that affect test results, including user competence assessment, risk assessment of testing kits and the environment in which they will be used, ease-of-use issues and the prevention of errors. We have previously described the care home workstream of the COVID-19 National Diagnostic Research and Evaluation Platform in Care Homes (CONDOR-CH), which is a service evaluation project to evaluate novel point of care testing technologies for COVID-19 designed to overcome shortcomings of currently used technologies [13]. The study reported here considers the use in care homes of a microfluidic immunofluorescence assay for the direct and qualitative detection of nucleocapsid protein antigen from SARS-CoV-2 (hereafter referred as LumiraDx). The COVID-19 testing strip for LumiraDx is designed to be used with samples collected from the anterior nares or nasopharyngeal site using a swab eluted into a vial of extraction buffer. A single drop of the specimen in extraction buffer is added to the test strip from a vial dropper cap. LumiraDx enacts the test protocol using dried reagents within the test strip. The result is determined from the amount of fluorescence detected within the measurement zone of the test strip. Analyte concentration in the specimen is proportional to the fluorescence detected. Results are displayed on the instrument’s touchscreen within 12 minutes of sample administration. Test strips are single use and disposed of post-assay. The system has been trialled and then subsequently deployed in different hospitals across England to inform decision making upon hospital admission.

This study aimed to:

- To evaluate diagnostic agreement, usability and biosafety risks of an automated antigen test.
- To generate risk minimisation and implementation strategies to enable test adoption in care homes.

## Methods

We conducted a mixed-methods (quantitative/qualitative) [14] evaluation with an agreement study of LumiraDx with qRT-PCR in care homes combined with a qualitative explorations of risks in use and to mitigate biosafety issues using unmoderated remote usability observation.

Homes were recruited using a national care home WhatsApp COVID-19 peer support group [15], public-facing social media and two national care home organisations: Care England and the National Care Forum. It includes nursing and residential homes; corporate chain, independent and third-sector providers; with between 20-350 residents per home. From this sampling frame we purposively selected four care homes (Appendix I, Table A1): we selected two homes with nursing and two without nursing (also known as nursing and residential homes respectively), The homes selected were in two regions of the UK, including two independent care homes and two from small chains of ownership. The sample was designed to maximise potential differences in staff training, and organisational configuration that might impact on implementation of a point-of-care test. The homes selected had previously been involved with the evaluation of a point-of-care Polymerase Chain Reaction (POC-PCR) test [13]. Their experience of deploying other POCTs was felt to provide useful contextual information for understanding LumiraDx use.

A range of participants were chosen to understand workflows around staff and resident routine testing. This sample enabled us to identify possible errors arising from repeated use of the machine, by different staff members, on different days, under different circumstances.

### Biosafety Issues

There are some challenges associated with deploying LumiraDx in care homes, which were important to understand and mitigate against:

1. Swabbing and eluting specimens into buffer is undertaken in bedrooms that are remote from the LumiraDx testing equipment raising potential biosafety issues (e.g.: from handling COVID-19 positive samples at the bedside, during transfer of the specimen, or upon arrival at the machine) due to risks of sample spillage of a potentially infected specimen.
2. As opposed to standard sample collection techniques for qRT-PCR tests, the swab must be swirled, squeezed and removed from the buffer, and then correctly disposed of in a biohazard bin. This procedure may expose staff and residents to risks (Fig. 1).

**Fig. 1.**
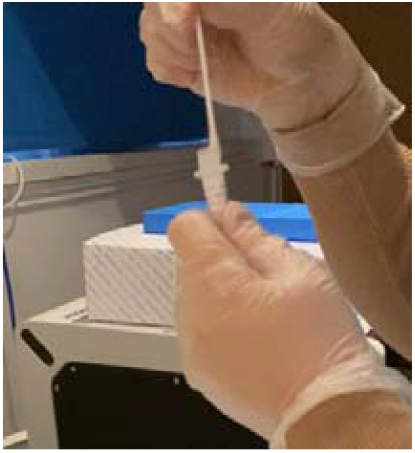
The operator removes the patient swab while squeezing the middle of the extraction vial to remove the liquid from the swab. The swab is discarded in biohazard waste.
3. The extracted sample is applied onto the test strip by gently pressing the sides of the extraction vial until one whole drop is visible and this procedure may cause contamination of the device and testing area (Fig. 2).

**Fig. 2.**
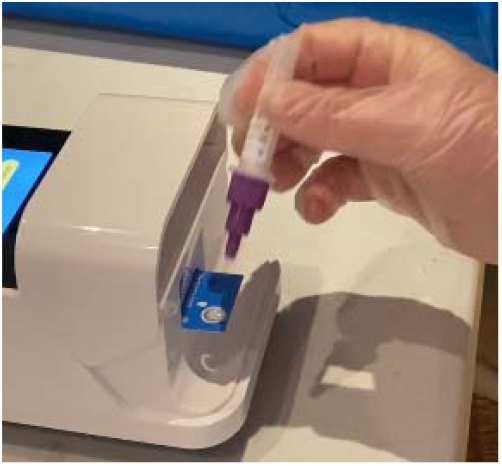
The operator applies the extracted sample from the extraction vial onto the sample application area of the inserted test strip.

### Simulation to evaluate biosafety issues

Four care homes took part in a trial with LumiraDx with no “live” nasal specimen. Each care homes allocated at least two participants for each site. Where applicable, participants were recruited to have mixed background and experience (i.e. staff involved in previous trials with point-of-care tests for SARS-CoV-2 and staff with no prior experience with point-of-care tests). A total of nine participants conducted the simulation test 5 times, independently, over one week. Staff were asked to follow a standardised operating procedure (SOP) prepared by laboratory staff taking account of perceived risks for eluting a swab, transferring the buffer solution, and running the LumiraDx test (see SOP; Appendix II).

Two randomly selected care homes (hereafter called care home A and care home B) were asked to conduct the simulation with GloGerm™ liquid, a mineral oil-based solution containing proprietary powder which fluoresces under ultraviolet light, which was used in place of buffer whilst following the SOP. After replacing LumiraDx buffer with GloGerm, we then asked them to examine their work and care areas under an ultraviolet light for signs of spillage or contamination. Verbalisation and interactions with the device were video recorded. Pictures of the testing area and device were taken at the end of each test. Following this, amendments were made to the SOP (Appendix III) and 10 further simulations [one volunteer in each care home (A and B) conducting five tests] were run to assess whether this had rectified the identified risks.

### Usability and use errors

Usability, potential sources of error, and ways of mitigating the risk (updated SOP) during routine test use with “live” specimens were the focus of semi-structured interviews with key stakeholders from four care homes. All interviews were undertaken remotely by a researcher in human factors (MM) at the conclusion of the trial. Interviews were semi-structured, lasted between 30 and 60 minutes, and were audio and video recorded with the interviewees’ permission. Interview schedules focused on manufacturer instructions for use, how LumiraDx might be integrated into the diagnostic pathway, the testing strategy and clinical decision making arising from positive and negative results. Interviewees from the four care homes were then prompted to explore potential usability issues such as clarity of test results, potential hazards and disposal procedures. Interviewees took part on a voluntary basis and did not receive compensation for their time.

Qualitative data were thematically analysed [16, 17]. Coding of the responses were performed by one researcher (MM) and codes were agreed following expert review with other human factors experts within the research team. Interviewees consented prior to the study and video interviews were recorded using conferencing platforms (MS Teams, Zoom.) Recordings were transcribed using AI transcription software (Otter.ai). Recording, transcription files and video materials were stored on a password-protected secure server. Enrolled participants were assigned a confidential identification number (IC) on consent, to be used on all corresponding data.

### Diagnostic agreement

The test was evaluated using specimens taken from both staff and residents as asymptomatic transmission from staff is an important factor in care home outbreaks [18]. We focused on routine staff testing; currently conducted three times per week, twice with lateral flow tests and once with send-off qRT-PCR. This high frequency, mixed regimen testing, generates significant workload for care home staff and makes that work more complicated [19]. We estimated that conducting 60 tests in each of the four care homes would allow enough repetition of the testing procedure to capture variability in practice between people and over time. We sought to conduct: 35 routine staff tests; 15 routine resident tests; and 10 tests for resident or staff who developed symptoms. Anterior nares swabs were taken by care home staff using the aforementioned SOP and adhering to standard testing procedures and kits recommended by the manufacturer (see Appendix II). All swab tests were taken by a member of the care home staff team. Staff did not self-swab. A paired nasopharyngeal swab for laboratory analysis was taken immediately after the LumiraDx swab using recommended UK Governmental guidance. Care home staff recorded LumiraDx test results using a results log, adding formal laboratory results when they became available. Unblinded but anonymised data were available to the research team. For the purpose of this study, LumiraDx was used in a dedicated testing area, fixed to a benchtop. Only formally trained staff members were permitted to use the machine.

No formal power calculation was undertaken. LumiraDx, in non-care home settings, has a reported sensitivity of 83.8% (95% CI: 76.4-89.2%) and specificity of 98.7% (95% CI 97.2-99.4%) with RT-PCR [20] as the reference method. Our objective was evaluating agreement between LumiraDx and qRT-PCR when the test was deployed in care home settings. All test results, including equivocal results and failures, were reported as per US Food and Drug Administration (FDA) guidance [21]. We calculated positive and negative agreement of LumiraDx with qRT-PCR results, (Cohen) Kappa, and the Brennan and Prediger statistic (equal to the prevalence and biased adjusted kappa (PABAK)) with associated 95% confidence intervals. Primary analysis was based on valid results for all tests stratified by symptomatic/asymptomatic participants. Calculations were carried out in Stata/SE 16.1 Sensitivity analysis to examine the impact of equivocal results and test failures was undertaken.

## Results

### Biosafety Simulations

Four main sources of contamination with GloGerm dye (Table 2) occurred. Table 2 shows UV LED images of operators after conducting the test with the GloGerm and mitigation strategies. These data along with interviews, led to the modification of the SOP (Appendix III) to improve glove, contaminated material and specimen handling at the bedside and the cleaning of surfaces after use. Simulation after modification showed no further evidence of contamination (Table 3).

**Table 1.**
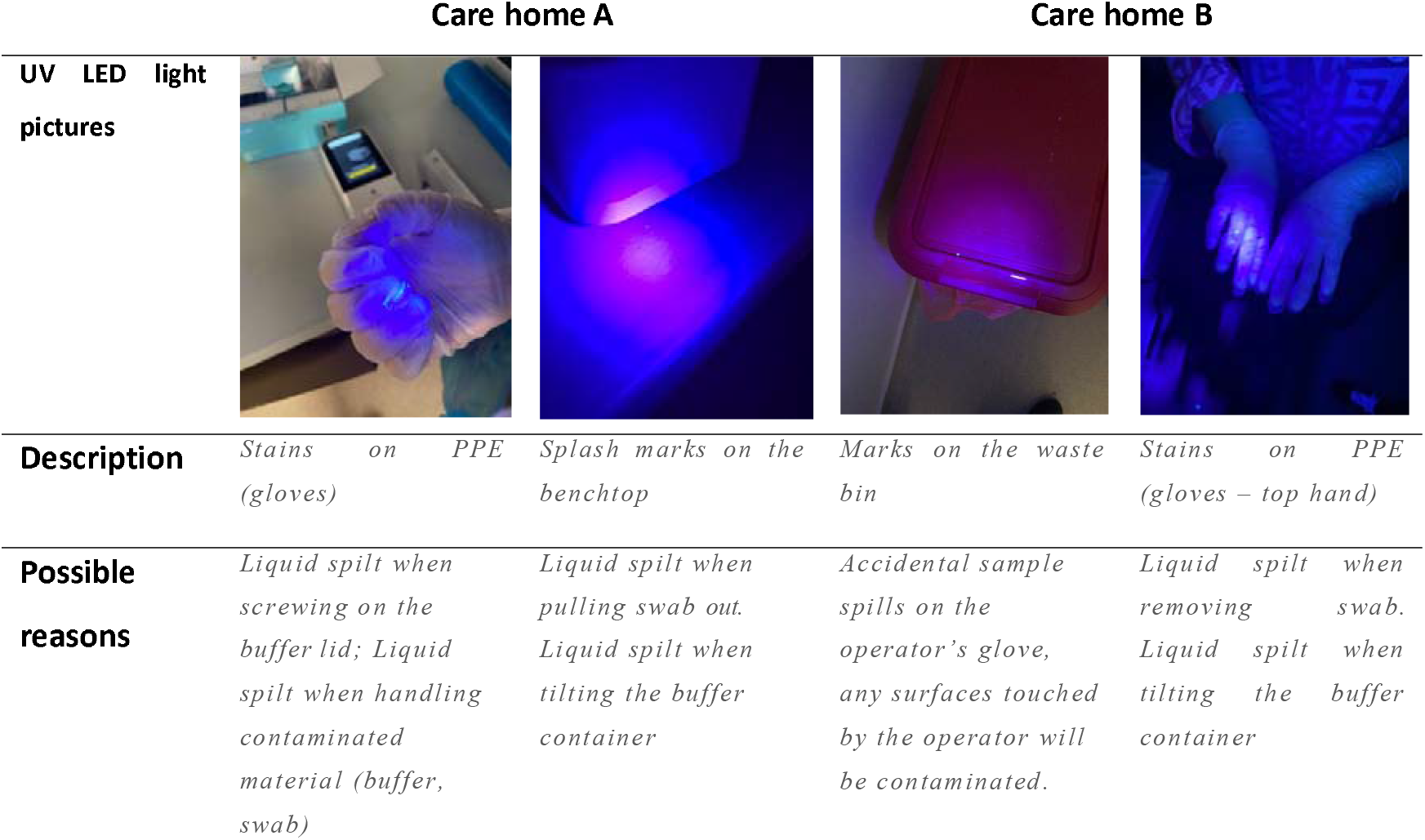
Biosafety issues identified using GloGerm dye during the simulation

|  | Care home A |  | Care home B |  |
| --- | --- | --- | --- | --- |
| UV LED light pictures |  |  |  |  |
| Description | Stains on PPE (gloves) | Splash marks on the benchtop | Marks on the waste bin | Stains on PPE (gloves – top hand) |
| Possible reasons | Liquid spilt when screwing on the buffer lid; Liquid spilt when handling contaminated material (buffer, swab) | Liquid spilt when pulling swab out. Liquid spilt when tilting the buffer container | Accidental sample spills on the operator's glove, any surfaces touched by the operator will be contaminated. | Liquid spilt when removing swab. Liquid spilt when tilting the buffer container |

**Table 2.** Full results from LumiraDx and Laboratory PCR

|  | Laboratory (qRT-PCR) |  |  |  |
| --- | --- | --- | --- | --- |
| LumiraDx | Negative | Positive | Uncertain/error | Total |
| Positive | 9 | 1 | 0 | 10 |
| Negative | 214 | 1 | 2 | 217 |
| Uncertain/error | 0 | 0 | 0 | 0 |
| **Total** | **223** | **2** | **2** | **227** |

Symptomatic cases
|  | Laboratory (qRT-PCR) |  |  |  |
| --- | --- | --- | --- | --- |
| LumiraDx | Negative | Positive | Uncertain/error | Total |
| Positive | 0 | 5 | 0 | 5 |
| Negative | 8 | 1 | 0 | 9 |
| Uncertain/error | 0 | 0 | 0 | 0 |
| **Total** | **8** | **6** | **0** | **14** |

**Table 3.**
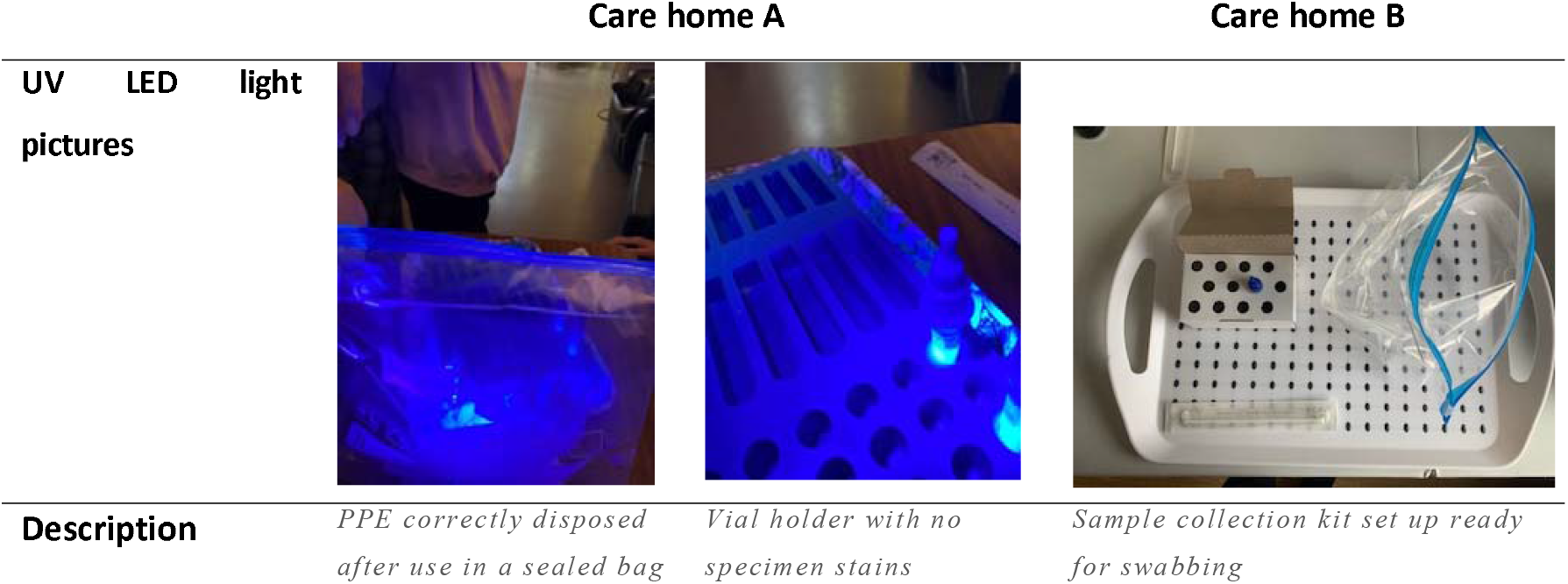
Dry-run with GloGerm dye adopting the amended SOP

|  | Care home A |  | Care home B |
| --- | --- | --- | --- |
| UV LED light pictures |  |  |  |
| Description | PPE correctly disposed after use in a sealed bag | Vial holder with no specimen stains | Sample collection kit set up ready for swabbing |

**Table 3.**
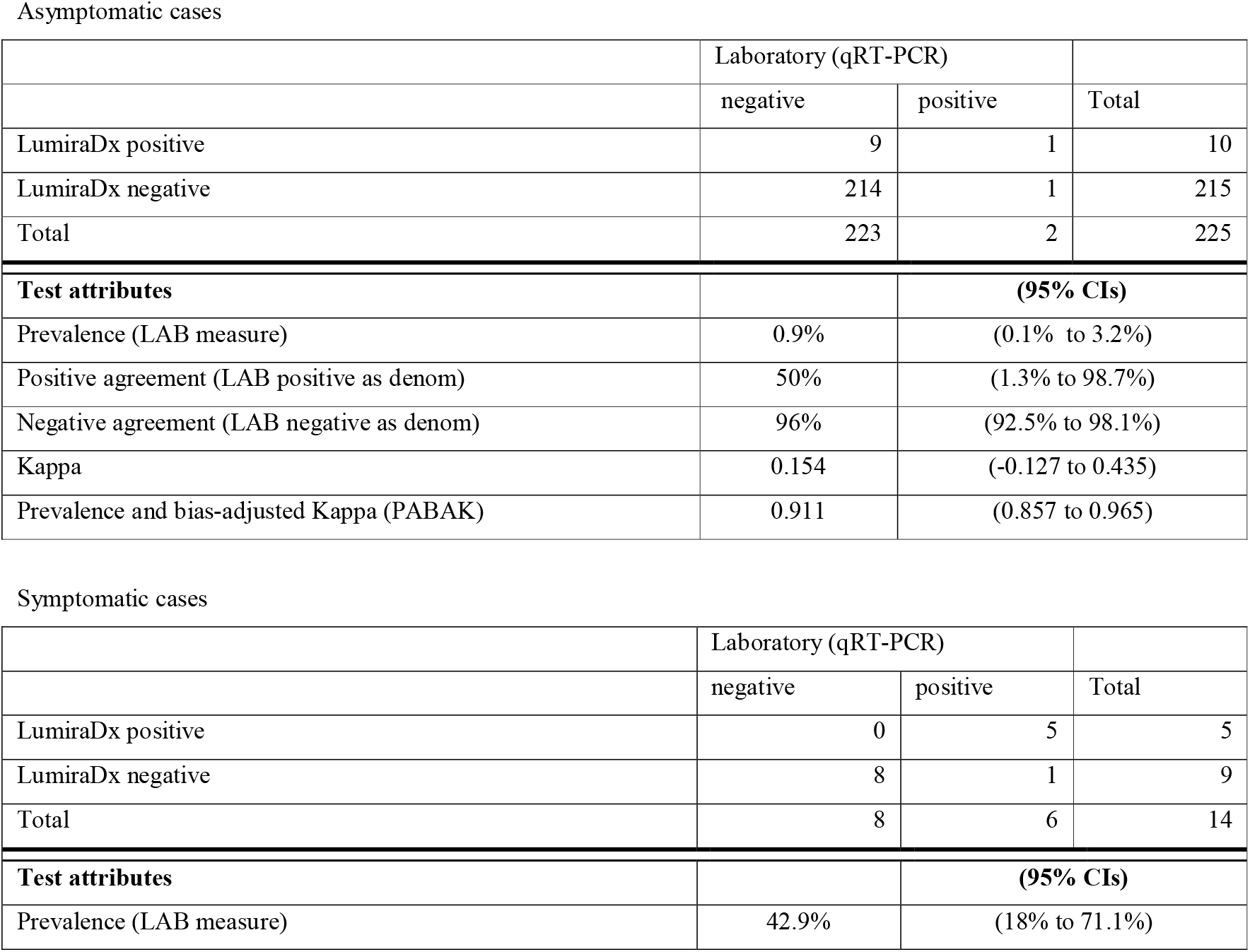
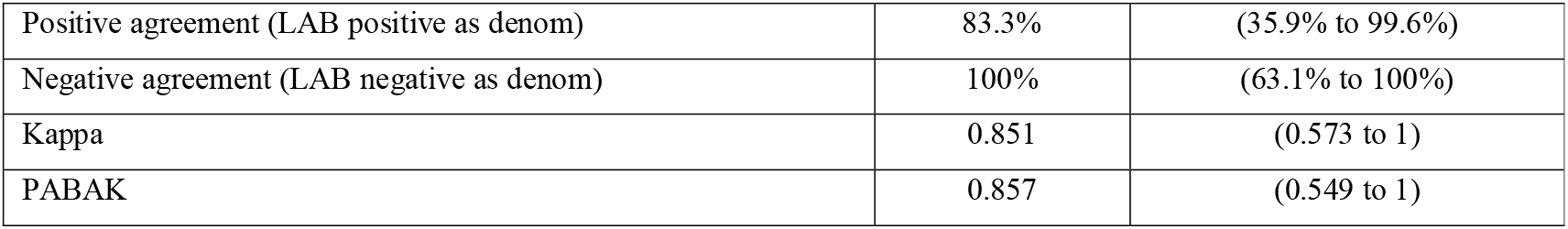
Agreement based on valid measures

|  | Laboratory (qRT-PCR) |  |  |
| --- | --- | --- | --- |
|  | negative | positive | Total |
| LumiraDx positive | 9 | 1 | 10 |
| LumiraDx negative | 214 | 1 | 215 |
| **Total** | **223** | **2** | **225** |
| **Test attributes** |  | **(95% CIs)** |  |
| Prevalence (LAB measure) | 0.9% | (0.1% to 3.2%) |  |
| Positive agreement (LAB positive as denom) | 50% | (1.3% to 98.7%) |  |
| Negative agreement (LAB negative as denom) | 96% | (92.5% to 98.1%) |  |
| Kappa | 0.154 | (-0.127 to 0.435) |  |
| Prevalence and bias-adjusted Kappa (PABAK) | 0.911 | (0.857 to 0.965) |  |

Symptomatic cases
|  | Laboratory (qRT-PCR) |  |  |
| --- | --- | --- | --- |
|  | negative | positive | Total |
| LumiraDx positive | 0 | 5 | 5 |
| LumiraDx negative | 8 | 1 | 9 |
| **Total** | **8** | **6** | **14** |
| **Test attributes** |  | **(95% CIs)** |  |
| Prevalence (LAB measure) | 42.9% | (18% to 71.1%) |  |

|  |  |  |
| --- | --- | --- |
| Positive agreement (LAB positive as denom) | 83.3% | (35.9% to 99.6%) |
| Negative agreement (LAB negative as denom) | 100% | (63.1% to 100%) |
| Kappa | 0.851 | (0.573 to 1) |
| PABAK | 0.857 | (0.549 to 1) |

### Usability

#### Interview findings focussed on three main areas

##### 1) Rapid test turnaround versus test throughput

Fast test turnaround (∼12 minutes) was valued. This was countered by an inability for staff members to handle more than one specimen at a time, and to carry on with their duties, especially if a large number of tests have to be conducted: *“The senior members of staff, the nursing staff, they are not going to be committed that level of time […] you’ve got to go back every 12 minutes. So, it’s taken up more of your time”*. (CH3)

Respondents identified specific use cases where LumiraDx was intuitively appealing: notably, sporadic testing such as testing of visitors at entrance to the home. Small changes to workflow were seen as potentially enabling the integration of LumiraDx into home routines, for example by staff testing themselves one at a time after arrival at work.

##### 2) Biosafety issues and appropriate staff user profile

Spillage was the main biosafety characteristic identifed. Participants recognised the mediating effects of safeguard measures and amended SOPs, but several staff felt testing was best undertaken by the more senior, and responsible, staff. Seniority was considered to be a function of time working in the home and/or could be denoted by a supervisor role. *“I don’t trust them as much [junior staff members], you know, to clean up after themselves…to use an expensive piece of equipment to not abuse that, to make sure that they’re writing the documentation down afterwards. So, for me, I would only be comfortable with somebody as a deputy manager or a manager level using the machine”*. (CH2)

LumiraDx is marketed as a bedside test for healthcare settings, but staff felt this to be inappropriate for a care home setting. This is because of the risk of spillage in areas with carpets and soft furnishing such as resident bedrooms. Further, it was seen as not appropriate in the communal parts of the home because the machine would be likely to attract attention from residents with cognitive impairment. Also, the machine made a noise during testing and the staff felt that residents would find this distressing, further limiting the proposed bedside use.

##### 3) Training materials

Respondents generally found training materials easy to follow but several reported that appropriate personal protective equipment (PPE), including visors, needed emphasising. Easily interpretable visual guides and a quick ‘prompt list’ mounted close to the machine would support operators recalling the operational steps, especially when use was infrequent (CH1).

### Testing agreement with laboratory RT-PCR

In total 241 tests were run. Tests were carried out on 227 asymptomatic participants (159 staff and 68 residents) and 14 symptomatic participants (5 staff and 9 residents) (Table 4; and Appendix IV, Table A2 stratified by staff/residents). Formal laboratory results were indeterminate for two specimens, both in asymptomatic participants, and so agreement analysis was conducted for 225 and 14 specimens. A Standard for Reporting of Diagnostic Accuracy Studies (STARD) Flow diagram is shown in Fig 3. The negative predictive value was 99.5% and 88.9% in asymptomatic and symptomatic participants, whilst positive predictive value was 10% and 100% in asymptomatic and symptomatic participants respectively. A Standard for Reporting of Diagnostic Accuracy Studies (STARD) Flow diagram is shown in Fig 3.

**Figure 3.**
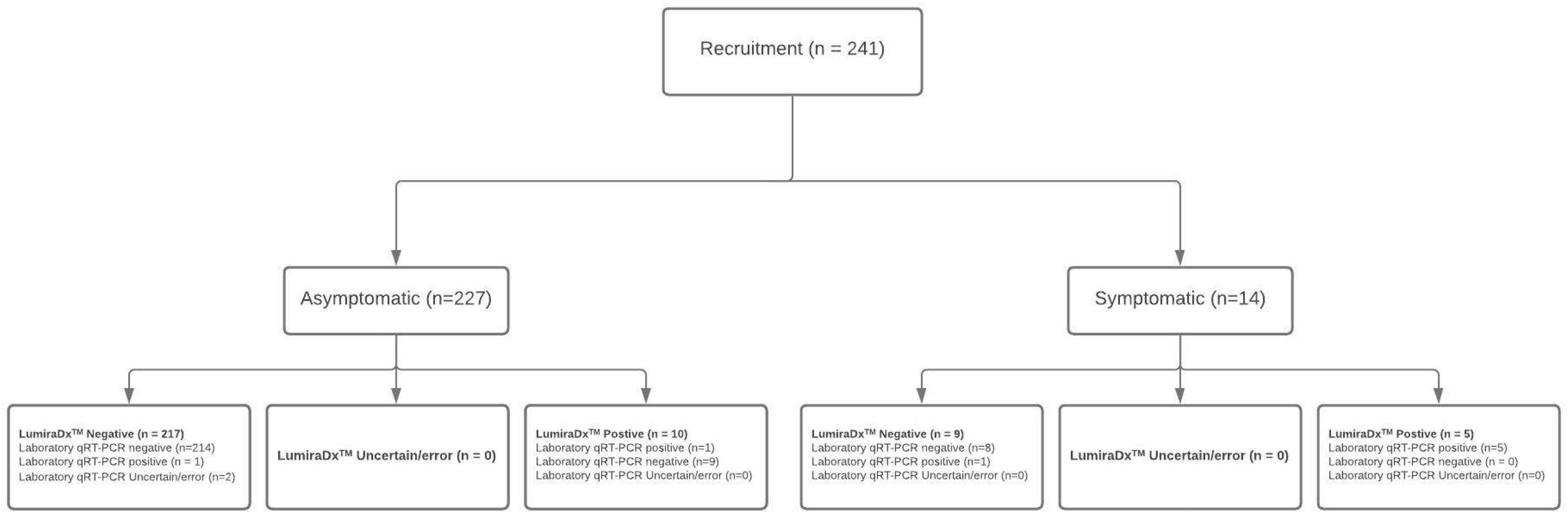
STARD diagram of samples acquired and test results

## Discussion

Our three main findings were that: i) usability observations are required to adjust SOPs for testing to take account of specific biosafety challenges arising from the care home context; ii) use (and thus usefulness) is potentially limited if not integrated into workflow; and iii) training materials need specific adaptations to match the information-literacy of care home staff.

As with other technologies we have evaluated in care homes [13], the utility of the LumiraDx test will depend on other factors including the care home built environment, staff competencies and workflow. The addition of LumiraDx to the range of tests that could be deployed in care homes, and the specific identification of a use case (e.g. testing of staff and visitors before entering the home), opens up a wider debate about what care homes require as we move into the next stage of the pandemic and beyond. Staff highlighted the potentially restrictive impact of using LumiraDx on workflow because of the existing workload of staff members. We also identified issues regarding how testing may influence other aspects of care delivery. Staffing ratios in care homes mean that staff sometimes struggle to meet routine care requirements even under normal circumstances [22]. The introduction of time-consuming, and at present statutorily mandated, POCTs risks distracting staff from routine caring tasks. This may compromise the ethos of care in within homes. The care sector should now consider whether these technologies might be purchased and retained in the sector, and how to plan staffing for their deployment without incurring excessive opportunity costs.

This was not a powered diagnostic accuracy study and the resulting negative and positive predictive values are a consequence of the low prevalence of positives in the group [23]. The good negative agreement suggests that LumiraDx may have some utility as a rule-out test. This could mean that it would work well in the context of a threshold test, which is the deployment suggested by care home staff. This would need to be further validated in a cohort study to estimate the test’s negative predictive value in this population. In this context, the false positive rate would mean the need for formal confirmation through formal RT-PCR testing for those who test positive.

The strengths of this study are that it provides a unique insight into the deployment of a novel POCT in care homes, a setting in which it has not previously been evaluated. The careful consideration of biosafety issues using GloGerm™ is, as far as we are aware, a first in this setting and could be replicated in future biosafety assessments in care homes. The in-context data on what is required if the technology is to be successfully adopted for care home deployment, could only have been acquired using the methodologies deployed. Study limitations include the small sample size. As such, the findings may not be representative of the 14,000 care homes across the UK. Further, it is recognised that the organisational factors identified in this sample of UK care homes may not be replicated internationally. Furthermore, we have limited knowledge of the physical properties of GloGerm™ (e.g. viscosity) as an appropriate surrogate for a human specimen. Nevertheless, the cautionary nature of our findings, about the need to evaluate new technologies in context in care homes, is likely to have wide application.

## Conclusions

LumiraDx was successfully and safely deployed in care homes following adaption of the standard operating procedure. This took account of the care home environment and staff training. LumiraDx increases the options for testing technologies that are available to care homes. Future candidate technologies for roll-out in this sector should be subject to similar rigorous mixed-methods evaluation. A consideration of the health economic impact and opportunity costs associated with roll-out of POCT in care homes is required.

## Supporting information

Appendix III

## Data Availability

Demographics of participants are included as supplementary mateiral

## Ethics

This project was approved as a service evaluation by Imperial College Healthcare NHS trust (ICHNT) – registration no. 471. The collation of data from routinely collected specimens taken as part of mandated patient care was determined using the Health Research Authority online toolkit to be a service evaluation.

## Acknowledgements

The authors would like to thank care home managers and staff members who took part in the study and members of the CONDOR platform for their comments: Mr Graham Prestwich, Ms Vale Tate, and Dr David Ashley Price.

