## Appendix I for "Point of Care Testing using rapid automated Antigen Testing for SARS-COV-2 in Care Homes – an exploratory safety, usability and diagnostic agreement evaluation"

Table A1. Care homes and staff operators included in the study

|  | **Test supervisor** | **Operators** | **Area** | **Nursing Home** | **Residential Home** | **TOT**  **number of beds** | **Identification Code (IC)** | **Previously involved in POCT**  **pilots** |
| --- | --- | --- | --- | --- | --- | --- | --- | --- |
| **CARE HOMEE A**  **(glow dye)** | Care Home  Manager |  | Nottinghamshire | x |  | 80+ | CH1 |  |
|  |  | Nursing  associate |  |  |  |  | Op1.1 | yes |
|  |  | House leader |  |  |  |  | Op1.2 | no |
| **CARE HOME B (glow dye)** | Managing  Director |  | Nottinghamshire |  | x | 140+ | CH2 |  |
|  |  | Project Improvement  Officer |  |  |  |  | Op2.1 | yes |
|  |  | Care home  manager |  |  |  |  | Op.2.2 | no |
| **CARE HOME C** (Not involved in glow dye trial) | Chief  Nursing Officer |  | Oxfordshire | x | x | 350+ | CH3 |  |
|  |  | Registered mental health nurse –  research  coordinator |  |  |  |  | Op.3.1 | yes |
|  |  | Pharmacy  technician |  |  |  |  | Op. 3.2 | no |
|  |  | Deputy  manager |  |  |  |  | Op. 3.3 | no |
| **CARE HOME C** (Not  involved in glow dye) | Managing  Director |  | Nottinghamshire | x |  | 50+ | CH4 |  |
|  |  | Training  coordinator |  |  |  |  | Op.4.1 | yes |
|  |  | Associate  nurse |  |  |  |  | Op.4.2 | no |
