## Appendix II for "Point of Care Testing using rapid automated Antigen Testing for SARS-COV-2 in Care Homes – an exploratory safety, usability and diagnostic agreement evaluation"

### **LumiraDx sars-cov-2 ag test: standard operating procedure**

This guide will provide you with step by step instructions on how to successfully use the LumiraDx to test individuals for COVID-19. Please ensure the steps are followed to enable the test to be completed in the correct manner for the best chance of an accurate result.

The LumiraDx SARS-CoV-2 Ag test is a rapid microfluidic immunofluorescence assay providing qualitative detection of nucleocapsid protein antigen to SARS-CoV-2.

**Health and safety:**

- When handling samples, full PPE must be worn **at all times** for your protection and protection of others. This includes gloves, gown, face masks and visors (if applicable)
- Once a swab is taken from a resident it must be tested within 5 hours.
- PPE should be discarded after taking the sample from the resident and then new PPE donned after entering the test room.
- Tests must be processed in a **separate, designated room** with all PPE worn and consumables disposed in waste streams provided.
- When testing is performed, only one person must be allowed into the room at any time
- Testing room should be **secured at all times** with the door kept closed
- Ensure all surfaces are cleaned down thoroughly with a disinfectant suitable for removing SARS-CoV-2.

**Training:**

Only members of staff that are trained to use the LumiraDx will be able to carry out testing.

**First use only:**

- Upon arrival of the instrument, plug into mains supply
- Ensure you are able to safely access the front of the machine to load/unload test strips and samples
- Once the instrument is plugged in and switched on, the homescreen with prompt for password

**For testing**

- Ensure the LumiraDx instrument is stationary on a flat surface
- Log in to the instrument using the log in details for USER found on the back of the bench top quick reference guide
- Select Result
- You will be prompted to add a strip lot into the instrument
- Lift the lid on the base of the instrument
- Follow the on screen instruction to push the strip into the exposed slot
- For a new lot number, the screen will state that the lot number of strip is not recognised
- You will be asked to register strip lot

#### **Registering a strip lot**

1. Tap the side of the strip box with the (((.))) facing the icon on the side of the analyser
2. The device will recognise and accept the strip lot number

#### **Running a Control**

1. Login to the instrument with used password
2. Select quality control on the screen
3. Open the box of controls, take out the in-use controls (red top negative, blue top positive
4. On the screen select ‘quality control test’
5. The screen will prompt you to add the strip
6. Remove strip from foil pouch and place into analyser
7. Using the micro pastette provided, take out some of the negative control
8. Add a drop onto the sample well (circle) on the strip lot
9. Close the lid and the instrument and wait a few seconds for the sample to detect
10. You will hear a beep and the assay will run
11. Repeat steps 6-9 for the positive control. Use a new strip lot

#### **Swab Sample collection and Sample Extraction**

You will need:

- Swab
- Extraction buffer vial
- Dropper lid

1. Wearing full PPE, obtain a nasal swab from each resident that requires a COVID test.
2. From the swab pack, take out the swab, an extraction buffer vial and the dropper lid
3. Swab the patient as per the diagram:
4. Remove the foil seal from the extraction buffer vial
5. Place the swab directly into the extraction buffer soaking the swab in the buffer for 10 seconds
6. Swirl the swab against the side of the vial 5 times
7. Remove the swab and dispose into a biological hazardous waste bin
8. Securely screw the dropper lid onto the vial ensuring a white filter can be seen in the lid
9. The sample must be kept upright and sent to the laboratory to be tested within 5 hours of collection

#### **Sample Testing**

3. Remove PPE and don fresh PPE on entry to the testing room.
4. The sample must be inverted 5 times to ensure proper mixing of buffer solution.
5. Log in to the instrument using the log in details for USER found on the back of the bench top quick reference guide
6. Select *Patient Test* on the touchscreen
7. Enter resident details
8. Remove a strip from its foil pouch holding the blue band at the bottom of the strip
9. Open the door of the instrument
10. Slot the strip into position until you feel resistance, the sample well (circle) must remain exposed
11. Ensure the vial of buffer is mixed well
12. Using the dropper, squeeze one drop of the buffer onto the sample well on the strip
13. Wait a few seconds for the sample to be detected
14. You will hear a beep and be prompted to close the door
15. Close the door and wait for the assay to run - *do not discard the buffer vial*
16. **DO NOT** knock the instrument while it is running this will invalidate the test
17. **If a test is invalid and requires a repeat, use a new strip lot but the same buffer vial**

**Results**

The test will run for 11-12minutes minutes, the screen will display a countdown to test completion.

Select *Finish* in the top righthand corner of the instrument and follow the instructions for removal of the test strip.

**Take care to keep the test strip flat when removing from the instrument so the sample does not run off.**

The used test strip must be disposed of into clinical waste.

#### **Quality Assurance procedures**

Positive and Negative controls are provided by manufacturer

Controls must be kept refrigerated at 2-8 degrees

When in use, ensure to insert lot number found on the box into the instrument on the touch screen

#### **Biological reference intervals, clinical decision values and critical alerts/values**

Ensure nasal swabs only are used to collect patient samples
