## Appendix III for "Point of Care Testing using rapid automated Antigen Testing for SARS-COV-2 in Care Homes – an exploratory safety, usability and diagnostic agreement evaluation"

### **LumiraDx sars-cov-2 ag test: AMENDED standard operating procedure (care homes)**

**Disclaimer:** This is a confidential document. The Standard Operating Procedures (SOP) in this document are under evaluation and should be used under the supervision of researchers at CONDOR.

Please refer to SOP provided by LumiraDx for tasks not included in this document.

**Test preparation (testing area)**

1. Prepare the exact number of extraction buffer vials to be used.
2. Label them with residents’ unique identifier

**In residents’ bedrooms**

You will need:

- A wipeable plastic tray
- A single-use sealable yellow clinical waste bag
- Alcohol cleaning wipes.

1. **Prepare the tray with the following:**
   a. **alcohol cleaning wipes (removed from packaging),**
   b. **sealable clinical waste bag widely open,**
   c. **and a vial holder (the cardboard holder provided by the manufacturer will suffice).**
2. Place the swab directly into the extraction buffer soaking the swab in the buffer for 10 seconds.
3. Swirl the swab against the side of the vial 5 times.
4. Remove the swab and dispose into the single-use clinical waste bag (Please be sure the swab does not touch the exterior of the sealable bag. The bag should be prepared on the tray before swabbing starts, as per point no. 1.b).
5. Securely screw the **blue lid** onto the vial.
6. Place the buffer tube down in the plastic tray.
7. The sample must be kept upright (please use carboard holder provided by the manufacturer) – test should happen within 5 hours of collection.
8. Wipe the plastic tray and exterior of the buffer tube with the alcohol wipe, before disposing of the alcohol wipe and gloves into the single-use clinical waste bag **(please DO NOT touch the alcohol wipe packaging. Wipes should be prepared before swabbing starts, as specified in point no. 1.a).**
9. Seal the bag.
10. Dispose of your other PPE and wash your hands as usual when leaving the room.
11. **Please DO NOT touch anything else in bedrooms (e.g. sharpies, door handle, etc…) before removing PPE used for sample collection.**
12. Go to the LumiraDx testing room where you commence the next step of the process.

**Sample testing (testing area)**

1. Remove PPE and don fresh PPE on entry to the testing room.
2. The sample must be inverted 5 times to ensure proper mixing of the buffer solution.
3. Replace the blue lid of extraction buffer vial with the dropper lid provided
4. Proceed with the standard testing procedure
5. **Pedal bin to be used to dispose of clinical waste**
6. **Wipe down the bench and the device in between uses with alcohol-based wipes.**
