## Appendix IV for "Point of Care Testing using rapid automated Antigen Testing for SARS-COV-2 in Care Homes – an exploratory safety, usability and diagnostic agreement evaluation"

Table A2. Full results from LumiraDx and Laboratory RT-PCR stratified by Staff vs. Residents

Asymptomatic

| Staff | Laboratory (qRT-PCR) | | |  |
| --- | --- | --- | --- | --- |
| LumiraDx | Negative | Positive | Uncertain/error | Total |
| Negative | 154 | 0 | 1 | 155 |
| Positive | 4 | 0 | 0 | 4 |
| Uncertain/error | 0 | 0 | 0 | 0 |
| Total | 158 | 0 | 1 | 159 |

| Resident | Laboratory (qRT-PCR) | | |  |
| --- | --- | --- | --- | --- |
| LumiraDx | Negative | Positive | Uncertain/error | Total |
| Negative | 60 | 1 | 1 | 62 |
| Positive | 5 | 1 | 0 | 6 |
| Uncertain/error | 0 | 0 | 0 | 0 |
| Total | 65 | 2 | 1 | 68 |

Symptomatic

| Staff | Laboratory (qRT-PCR) | | |  |
| --- | --- | --- | --- | --- |
| LumiraDx | Negative | Positive | Uncertain/error | Total |
| Negative | 3 | 0 | 0 | 3 |
| Positive | 0 | 2 | 0 | 2 |
| Uncertain/error | 0 | 0 | 0 | 0 |
| Total | 3 | 2 | 0 | 5 |

| Resident | Laboratory (qRT-PCR) | | |  |
| --- | --- | --- | --- | --- |
| LumiraDx | Negative | Positive | Uncertain/error | Total |
| Negative | 5 | 1 | 0 | 6 |
| Positive | 0 | 3 | 0 | 3 |
| Uncertain/error | 0 | 0 | 0 | 0 |
| Total | 5 | 4 | 0 | 9 |
